# Detection of SARS-CoV-2 in air and on surfaces in rooms of infected nursing home residents

**DOI:** 10.1101/2022.02.16.22271053

**Authors:** K.J. Linde, I.M. Wouters, J.A.J.W. Kluytmans, M.F.Q. Kluytmans-van den Bergh, S.D. Pas, C.H. GeurtsvanKessel, M.P.G. Koopmans, M. Meier, P. Meijer, C.R. Raben, J. Spithoven, M.H.G. Tersteeg-Zijderveld, D.J.J. Heederik, W. Dohmen, COCON consortium

## Abstract

There is an ongoing debate on airborne transmission of Severe Acute Respiratory Syndrome Coronavirus 2 (SARS-CoV-2) as a risk factor for infection. In this study, the level of SARS-CoV-2 in air and on surfaces of SARS-CoV-2 infected nursing home residents was assessed to gain insight in potential transmission routes.

During outbreaks, air samples were collected using three different active and one passive air sampling technique in rooms of infected patients. Oropharyngeal swabs (OPS) of the residents and dry surface swabs were collected. Additionally, longitudinal passive air samples were collected during a period of 4 months in common areas of the wards. Presence of SARS-CoV-2 RNA was determined using RT-qPCR, targeting the RdRp- and E-genes. OPS, samples of two active air samplers and surface swabs with Ct value ≤35 were tested for the presence of infectious virus by cell culture. In total, 360 air and 319 surface samples from patient rooms and common areas were collected. In rooms of 10 residents with detected SARS-CoV-2 RNA in OPS, SARS-CoV-2 RNA was detected in 93 of 184 collected environmental samples (50.5%) (lowest Ct 29,5), substantially more than in the rooms of residents with negative OPS on the day of environmental sampling (n=2) (3.6%). SARS-CoV-2 RNA was most frequently present in the larger particle size fractions (>4 μm 60% (6/10); 1-4 μm 50% (5/10); <1 μm 20% (2/10)) (Fischer exact test p=0.076). The highest proportion of RNA-positive air samples on room level was found with a filtration-based sampler 80% (8/10) and the cyclone-based sampler 70% (7/10), and impingement-based sampler 50% (5/10). SARS-CoV-2 RNA was detected in ten out of twelve (83%) passive air samples in patient rooms. Both high-touch and low-touch surfaces contained SARS-CoV-2 genome in rooms of residents with positive OPS (high 38% (21/55); low 50% (22/44)). In one active air sample, infectious virus *in vitro* was detected.

In conclusion, SARS-CoV-2 is frequently detected in air and on surfaces in the immediate surroundings of room-isolated COVID-19 patients, providing evidence of environmental contamination. The environmental contamination of SARS-CoV-2 and infectious aerosols confirm the potential for transmission via air up to several meters.

## Introduction

The ongoing pandemic, caused by Severe Acute Respiratory Syndrome Coronavirus 2 (SARS-CoV-2), continues to constitute a public health emergency of international concern. There is consensus on the role of direct contact transmission and airborne transmission at short distances (up to several meters) through large droplets. There is an ongoing debate on transmission through fomites and airborne transmission at larger distances (up to more than several meters) as a risk factor for subsequent infection ^1,2^. The relative importance of this mode of transmission as driver of the pandemic is unknown. Several modes of transmission through the environment as a possible risk of infection of SARS-CoV-2 is considered important for groups at high risk ^3^.

Previous studies investigating SARS-CoV-2 air concentrations in healthcare facilities showed contradictory results. In a limited number of studies in hospital settings, SARS-CoV-2 has been detected in air in proximity (2-5m) of COVID-19 patients ^1,4–8^. Other studies did not find evidence of SARS-CoV-2 in air ^9–12^. However, the comparability of studies is limited due to differences in sampling methods, sampling duration and distance to infected persons. Oropharyngeal swabs were not consistently collected from infected persons for confirmation of infection and the actual level SARS-CoV-2 shedding in addition to the collection of air samples. Infectiousness of SARS-CoV-2 detected in air was not investigated in most studies ^4–7^ or could not be shown ^13,14^. Infectivity and amount of shed virus have been reported to rapidly decline during the first week after illness onset ^15,16^. As viral RNA can persist and be shed for prolonged periods of time without being infectious, it is important to investigate the viability of virus in air to understand airborne transmission routes of the virus. Therefore, to successfully investigate modes of transmission of SARS-CoV-2, it seems crucial to investigate SARS-CoV-2 air concentrations in the first days following infection.

After the first pandemic wave in the Netherlands, nursing homes had introduced enhanced surveillance screening for SARS-CoV-2, which led to identification of new infections at an early stage ^17^. To determine airborne transmission risks from SARS-CoV-2 infected patients to their immediate surroundings, we measured SARS-CoV-2 in air and on surfaces in Dutch nursing home residencies as well as in rooms of SARS-CoV-2 isolated infected nursing home residents.

## Methods and materials

The study consisted of two arms: a series of environmental investigations during outbreaks and longitudinal air monitoring (Figure 1). Weekly SARS-CoV-2 infections were registered and notified in 28 nursing homes from Mijzo Care organisation in Noord-Brabant in the Netherlands. In case of two or more confirmed SARS-CoV-2 infections in residents within the same ward, an outbreak investigation was initiated, consisting of extensive environmental sample collection and SARS-CoV-2 testing of persons. In a subsample of 3 of the 28 nursing homes, longitudinal monitoring took place in the direct living environment. No medical ethical approval was needed for this study as evaluated by the Medical Research Ethics Committee of University Medical Centre Utrecht; a declaration of non-compliance with the scope of the Dutch Medical Research Involving Human Subject Act was obtained. The study was conducted in agreement with the European legislation on handling privacy-sensitive data.

**Figure 1:**
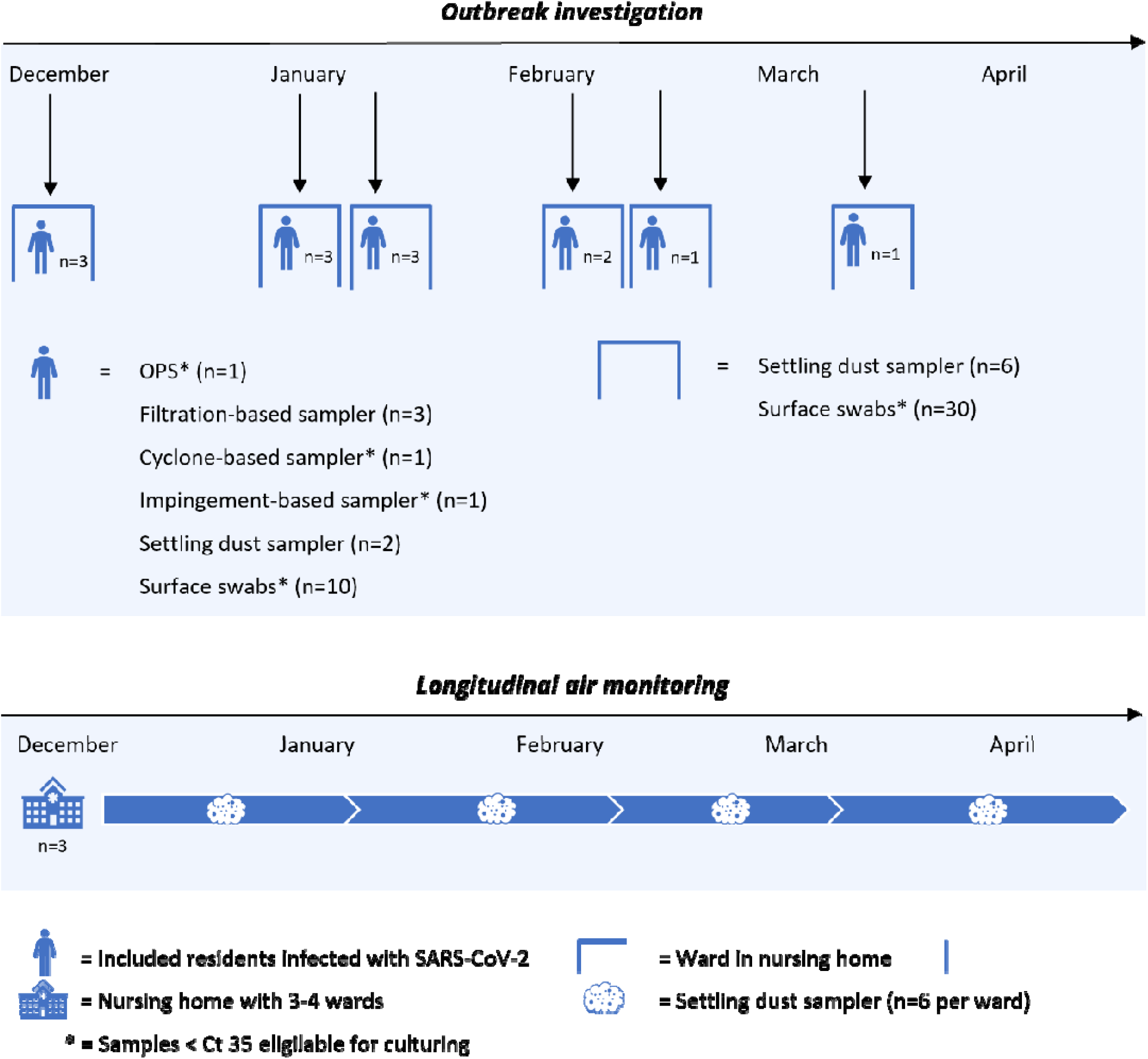
Design of the study. The study consisted of an outbreak investigation which was complemented by longitudinal air monitoring.

### Outbreak investigation

Residents of the nursing homes were tested for possible SARS-CoV-2 infection in case they experienced COVID-19 related symptoms. When one or more residents in a ward tested positive for SARS-CoV-2 infection, all other ward residents were screened for SARS-CoV-2 infection during surveillance rounds. Residents who tested positive for SARS-CoV-2 RNA were eligible for inclusion in the outbreak investigation within 8 days since the onset of symptoms or within 8 days since the first positive surveillance test result. Only patients in isolation from somatic wards were included. Oral informed consent was obtained from patients and/or from an authorised legal representative or family member.

#### Collection of air samples

Air samples were collected at three locations in the patient’s room: 1) near the head of the patients within approximately 0.5 meter of the patient, 2) near the feet of bedridden patients approximately 1.5 meters from the head or approximately 1.5 meters from mobile patients sitting in a chair, and 3) near the location often used by healthcare workers more than 2 meters away from the patient such as the sink, all positioned at 1.5m height. In every patient room, 6-hr inhalable dust samples were taken using a filtration-based technique at all three locations (Conical Inhalable dust Sampler (CIS), JS Holdings, UK). In addition, one 6-hr two-stage cyclone-based sample with filter back-up was positioned near the feet of the patient when bedridden or at 1.5 meters from the chair of the patient (NIOSH BC 251, kindly provided by Dr William G Lindsley, NIOSH CDC, Morgantown, USA), as well as a 1-hr impingement-based sampler positioned in proximity of the head of the patient (5ml BioSampler, SKC, UK) (See supplement Figure S1). During the 6-hour sample collection, mobile patients were allowed to move in the room. During the 1-hour impingement-based sample collection, they were asked to stay seated in their chair. The filtration-based sampler was equipped with a 37mm diameter 2.0μm pore-size Teflon filter (Pall incorporated, Ann Arbor, USA). The two-stage cyclone-based sampler allowed size-selective sampling and was equipped with two conical tubes (of 15 ml and 1.5 ml) which sample respectively particulates of 1-4μm and >4μm, and a back-up Teflon filter (37 mm diameter 2.0 μm pore-size Pall incorporated, Ann Arbor, USA) for particulates of <1μm when operated at a flow of 3.5L/min. The 15 ml and 1.5 ml conical tubes were filled with virus transport medium 1 (VTM-1; Erasmus Medical Center (EMC), Rotterdam, The Netherlands) during sampling and Opti-MeM™ (Gibco, UK) was added immediately after collection (see supplementary methods for more details and composition of media). Adding VTM-1 is a modification of the standard operating procedure for this sampler with the aim to enhance culturability of the virus. The impingement-based sampler contained VTM-1 during sampling, and after completion of sampling, Opti-MeM™ was added as well.

Airborne settling dust was sampled using Electrostatic Dust Collectors (EDCs) ^18^, which were placed in each included patient room and the corresponding hallway, common living room, and nurse office of the ward. EDCs were placed in holders pinned to the ceiling in the middle of the space, approximately 30cm underneath the ceiling or on top of a cabinet. EDCs were collected after 2-4 weeks of sampling, dependent on the timing of extensive cleaning of the room.

#### Collection of surface samples

High- and low-touch surface samples were collected using dry surface swabs (Medical Wire Dry Swabs, MW730, Corsham, UK) as described previously ^19,20^. A total of ten samples were taken in each patient room, and in the corresponding hallway, common living room, and nurse office of the ward. Disposable plastic grids of 10 cm^2^ were used to standardise collection of surface swabs. Swabs were placed in viral transport medium 2 (VTM-2; Erasmus Medical Center (EMC), Rotterdam, The Netherlands) directly after collection (see supplementary methods for the composition of media).

Field blank samples were collected every other outbreak sampling day for each air sampling technique and every outbreak sampling day for surface swab sampling. See supplementary methods for details on sample collection and laboratory methods.

#### Patient characteristics

Patient characteristics were obtained: gender, year of birth, date of symptom onset, symptoms, date of SARS-CoV-2 test, SARS-CoV-2 test results, COVID-19 treatment such as oxygen therapy and mobility. An affirmative oropharyngeal swab (OPS) (Medical Wire Dry Swabs, 111598, Milano, Italy) was collected during the outbreak investigation and stored in a tube containing VTM-2.

### Longitudinal air monitoring

Three of the 28 nursing homes with at least three wards (somatic and/or geriatric) were selected for longitudinal air monitoring. In each selected nursing home, settling dust samples were collected repeatedly in three to four wards from December 2020 until May 2021. Per ward, 6 EDCs were placed in hallways, living rooms and nurse offices and renewed every four weeks for a period of four months. Incidence of SARS-CoV-2 infections in patients and staff members at the included wards was obtained in weekly reports.

### Laboratory analysis

All samples, except settling dust samples, were stored and send to the laboratory refrigerated at 4 °C directly after collection. At the laboratory, samples were stored at 4°C until further processing within 24-hours under biosafety laboratory (BSL)-2+ conditions ^21^. Filters were removed from the filter holder and transferred to a tube containing VTM-1. These and all other outbreak investigation samples were subsequently vortexed. Settling dust samples were transferred to tubes containing VTM-2 and tamped down, vortexed and soaked repeatedly for several minutes. For RT-qPCR analysis, an aliquot of VTM was mixed in a 1:1 dilution with MagNA Pure 96 External Lysis Buffer for each sample (Roche Diagnostics, Almere the Netherlands). Remaining VTM from cyclone-based samples, impingement-based samples, surface swabs and OPS were stored for culturing. All samples were stored frozen at -80 °C until further processing. More details are described in the supplementary methods.

#### Real time semi-quantitative reverse transcriptase polymerase chain reaction

VTM-lysis buffer samples were tested for presence of SARS-CoV-2 RNA using a SARS-CoV-2 RNA RT-qPCR, targeting the E gene and CoV-2 RdRp-gene of SARS-CoV-2 using the cobas® 6800/8800 Systems (Roche Diagnostics) ^22^. If both E-gene and RdRp-gene were detected with Cobas RT-qPCR, samples were classified as positive. In case of a discrepant result, i.e. only one of the two genes was detected, an in-house RT-qPCR assay was conducted for confirmation ^23^. In case of detection of SARS-CoV-2 RNA, the sample was classified as positive and in case of inconclusive or non-detection with in-house RT-qPCR, the final result was classified as inconclusive. Samples with non-detection of both genes were classified as negative.

#### Virus culture

OPS, cyclone-based samples, impingement-based samples, and surface swabs tested positive by RT-qPCR with RdRp Ct ≤35 were tested for infectious SARS-CoV-2, as described previously ^16^. Virus culture was performed in 24-wells plates seeded with Vero cells, clone 118. Samples were added to the wells, centrifuged, and inoculum was discarded. Virus culture medium was added, and samples were cultured at 37°C and 5% CO_2_ for seven days. If a virus-induced cytopathic effect (CPE) was observed, immunofluorescent detection of SARS-CoV-2 nucleocapsid protein was performed to confirm the presence of SARS-CoV-2.

### Data analysis

Data entry was carried out in Microsoft Access Version 16 2012. Descriptive statistics were obtained by R studio Version 1.4.1106 2021. Active air sample techniques were compared on room level. If one or more of the filtration-based samples in a room were positive, the outcome on room level was classified as positive. The same applied for the CDC-NIOSH cyclone-based samples on room level. Fisher’s exact test was used to compare the proportion of positive samples in association with particle size fractions, distance and location, and to compare air sampling techniques. Agreement between outcomes of filtration-based and cyclone-based samples collected at the same location was investigated through Cohen’s Kappa test statistics. A threshold of 0.05 was used for the p-value for statistical significance.

## Results

A total of 679 environmental samples were collected from five nursing home wards, including 101 air samples and 122 surface samples from the patient rooms and 259 air samples and 197 surface samples from common areas. In total, 13 patients were included for environmental sample collection during outbreak investigations. One patient withdrew from the study during sampling. Of the remaining 12 patients, two tested negative, and 10 tested positive in affirmative OPS collected on the day of environmental sample collection (Table 1). From one patient with negative OPS, only surface swabs were collected. For air samples, RdRp Ct-values ranged from 29.5 to 37.2 and from 30.2 to 37.8 in surface swab and from 19.8 to 34.7 in OPS. All field and laboratory blanks tested negative for viral SARS-CoV-2 RNA.

**Table 1:**
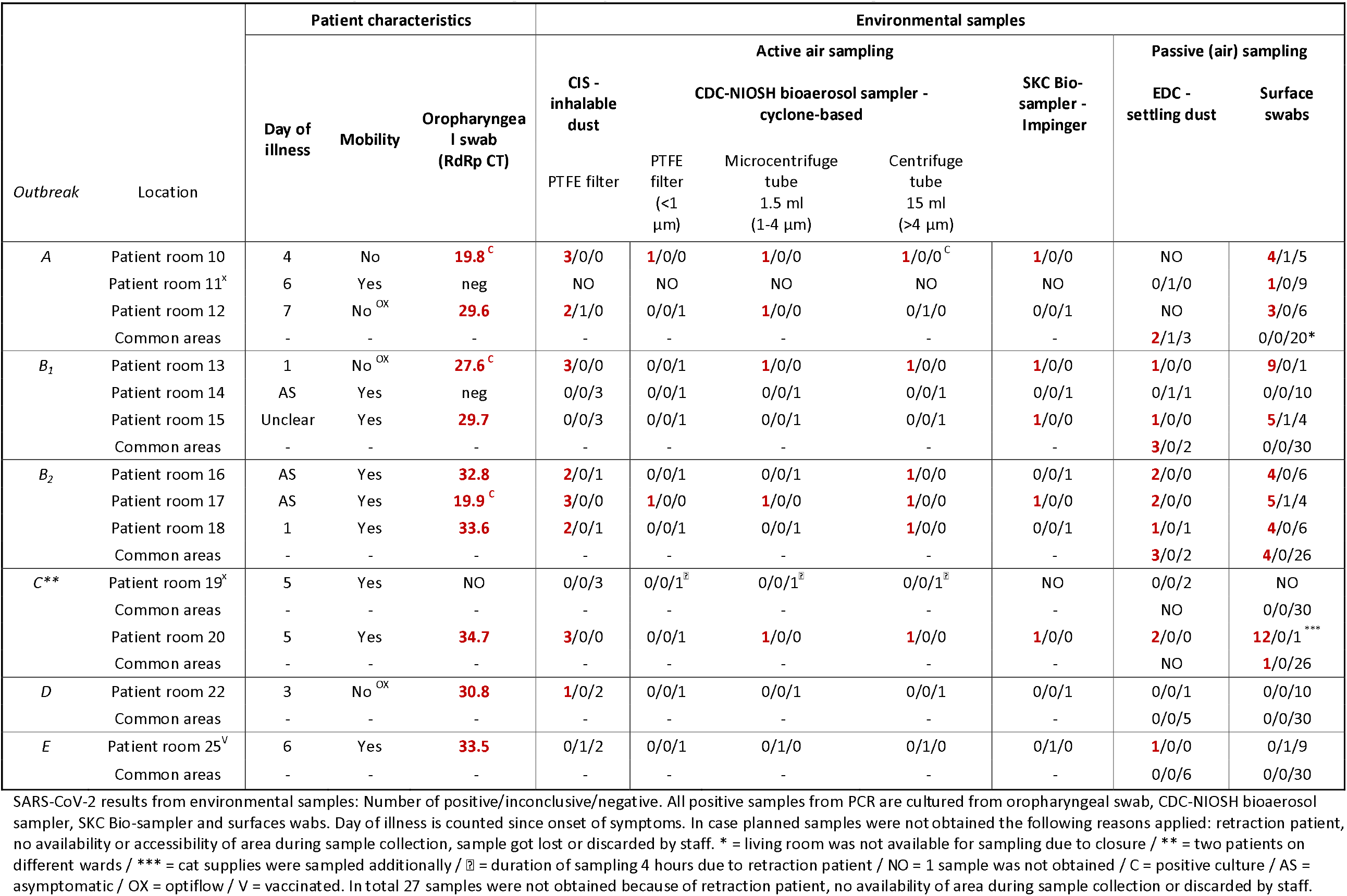
SARS-CoV-2 PCR in environmental samples in the surrounding of isolated patients and common areas in nursing homes.

### SARS-CoV-2 contamination in air

Of the 184 environmental samples collected in rooms of patients with positive OPS on the day of sampling, 50.5% were positive (93/184). From the two patients with negative OPS, only one of the samples tested positive (1/29), which appeared a surface swab (Table 1).

All four air sampling techniques detected SARS-CoV-2 RNA and showed high rates of positive samples in the rooms of patients with positive OPS (Table 3). The highest proportion of positive active air samples was found with the filtration-based sampler 80% (8/10) and CDC-NIOSH cyclone-based sampler (70% (7/10). The impingement-based sampler (50% (5/10)) showed a slightly lower proportion of positive samples, but the results were not statistically significant (Fisher-exact test p-value=0.69). The cyclone-based samples sampled approximately 1.26 m^3^ of air, the filtration-based samples 1.26 m^3^ and the impingement-based 0.75 m^3^. Ten of the collected 12 settling dust samples from rooms were positive (83%). Filtration-based samples and cyclone-based samples collected side-by-side at the same distance from the patient were concordant in 8 out of 10 cases (moderate agreement (Cohen’s kappa coefficient kappa=0.5, p-value=0.197)) (Supplement Table S4).

**Table 2:**
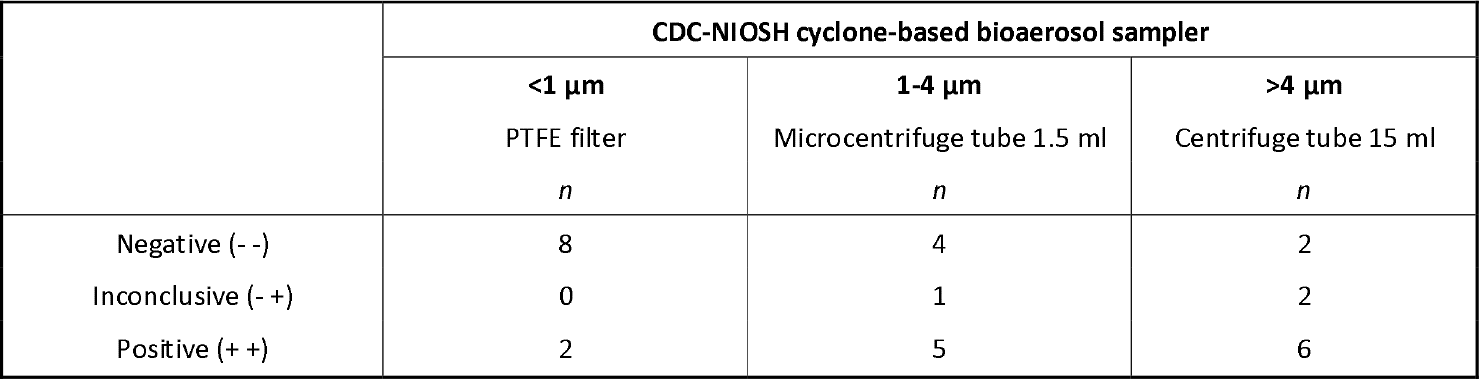
SARS-CoV-2 PCR results in size specific fractions obtained by cyclone-based air sampling in rooms of patients with positive oropharyngeal swab.

**Table 3:** SARS-CoV-2 results from three active and one passive air sampling technique used during the outbreak investigation from patients with positive oropharyngeal swab.

|  | CIS -<br>inhalable dust<br>*n* (%) | SKC Bio-sampler -<br>Impinger<br>*n* (%) | CDC-NIOSH cyclone-based bioaerosol *<br>*n* (%) | EDC -<br>settling dust<br>*n* (%) |
| --- | --- | --- | --- | --- |
| Negative (- -) | 9 (30) | 4 (40) | 2 (20) | 2 (17) |
| Inconclusive (- +) | 2 (7) | 1 (10) | 1 (10) | 0 (0) |
| Positive (+ +) | 19 (63) | 5 (50) | 7 (70) | 10 (83) |

\* = if one of the fractions of the cyclone-based sample detected SARS-CoV-2, the overall parameter is classified positive.

SARS-CoV-2 was detected at all distances from the patient. No clear trend was seen in numbers of positive samples with distance from the patient in filtration-based air samples (>1.5m 50% (6/12); ≤1.5m 67% (10/15)) (Fisher-exact test, p-value=0.4175) (Supplement Table S3).

In all particle size-specific fractions (>4 μm 60% (6/10); 1-4 μm 50% (5/10); <1 μm 20% (2/10) SARS-CoV-2 RNA was detected (Table 2). However, inconclusive and positive results were more frequently present in the largest particle size fraction, followed by the intermediate size fraction. These differences in distribution of size categories was borderline statistically significant (Fischer exact test p-value=0.076).

### High- and low-touch surface swabs

The proportion of positive surface samples was much higher in rooms from patients with positive OPS compared to rooms with negative patients (43% (43/99) versus 0,5% (1/20)) (see Supplement Table S5). SARS-CoV-2 RNA was detected slightly more frequently in surface swabs from low-touch surfaces than from high-touch surfaces (low 50% (22/44); high 38% (21/55)) (Fisher’s exact test p-value=0.18). Only 5 of the 197 surface samples collected in common areas were positive for SARS-CoV-2; four low and one high-touch sample (Supplement Table S6).

### Virus culture

Among the 78 positive OPS, cyclone-based samples, impingement-based samples, surface swab samples, 44 had a RdRp Ct-value ≤35 and were further investigated by *in vitro* virus culture. This selection contained four impingement-based samples, three cyclone-based samples fraction size >4 μm, three cyclone-based samples fraction size 1-4 μm, 26 surface swabs and eight OPS collected in nine patient rooms. The impingement-based samples and cyclone-based samples were collected in four patient rooms. Cytopathic effects were observed in three OPS and one active air sample and were confirmed by immunofluorescent staining. The active air sample from the CDC-NIOSH sampler (>4µm size fraction) had the lowest Ct-value of all environmental samples (29.5) and was derived from the room of the patient with the lowest OPS Ct-value (19.82).

### Whole genome sequencing (WGS)

In total, nine samples with RdRp Ct-values ranging from 19.8 to 30.2 were selected for SARS-CoV-2 whole-genome sequencing, of which six OPS, one cyclone-based sample, one filtration-based sample and one surface swab. From five OPS samples, >90% of the reference was covered and uploaded in GISAID. All variants were B.1.221, a known variant, circulating in the Netherlands at the time of the study. Samples collected at the same location were closely genetically related. During the data collection from December 2020 until May 2021, B.1.1.7, also known as the Alpha variant, became the dominant SARS-CoV-2 circulating variant in the Netherlands ^24^. The sequences have been registered in GISAID (www.gisaid.org; Accession ID EPI_ISL_2259112, EPI_ISL_2259136, EPI_ISL_2259188). See supplementary for more details and acknowledgement table.

### Longitudinal air monitoring

Only seven of the 259 settling dust samples collected repeatedly in three wards were positive (2.7%). All samples were collected in common areas in nursing homes where SARS-CoV-2 infections had been reported among residents (Supplement Table S7). The low rate corroborates with the incidence of infections in patients and healthcare workers, which rapidly decreased during the study (Table S7). No viral RNA was detected in wards without registered SARS-CoV-2 infected patients and/or healthcare workers shortly before or during sampling.

## Discussion

In this study, comprising 679 environmental samples, SARS-CoV-2 was frequently detected in air and on surfaces in the immediate surroundings of COVID-19 patients, providing evidence of virus shedding to the environment through air by infected persons. SARS-CoV-2 was detected more frequently in the particle size fraction 1-4μm (respirable fraction) and particulates >4μm as compared to <1μm. Airborne particulates might be infectious, as illustrated by the fact that we were able to replicate virus from an active air sample. Our results support the role of airborne transmission of SARS-CoV-2, which in turn is a risk factor for subsequent infection.

SARS-CoV-2 RNA was detected in all types of air samples and on high and low-touch surfaces in the surrounding of patients with a positive OPS. No SARS-CoV-2 RNA was detected in air or the immediate surroundings of patients who tested negative. The number of positive environmental samples in this study was high compared to other studies ^1,25^. Although the study size is small to modest, environmental sampling was performed extensively around patients in early phase of infection, assuming active shedding of SARS-CoV-2. Previously, Van Beek et al., established a shedding curve using data from 223 persons testing SARS-CoV-2 in a drive-through test station, showing that viral loads were highest within eight days post onset of symptoms ^15^. Moreover, Van Kampen et al., reported that infectious virus shedding also occurred mainly within the first eight days post-onset, based on data from 129 hospitalised patients with repeated measurements ^16^. Therefore, our study’s timing of environmental measurements has likely contributed to the high detection rate in environmental samples. This is in agreement with a study from Chia et al. only detecting SARS-CoV-2 in air or the immediate surroundings of two patients infected less than eight days compared to no detection of SARS-CoV-2 in the air of another patient nine days post-infection ^4^. Several other studies were not able to detect SARS-CoV-2 in air in the surrounding of patients more than eight days after post-onset of symptoms ^4,25,26^. Also, studies in non-healthcare settings without patients in the early phase of infection, such as in secondary schools in the aftermath of SARS-CoV-2 associated outbreaks, were not able the measure detectable levels of SARS-CoV-2 RNA (personal communication Dr. Wouters, 2021). However, SARS-CoV-2 was detected in great abundance in surroundings from infectious minks in acute phase in mink farms ^19^. These observations emphasize that timing of sampling in the direct environment of patients and other populations is of importance for detecting SARS-CoV-2.

Of the SARS-CoV-2 containing aerosols, 54% was in the size range <4µm and 46% in the size range of ≥4µm. When including samples with inconclusive qPCR test results, these figures hardly changed (50%-50%). Although the use of the NIOSH sampler was modified by adding VTM to the vials prior to sampling, which may theoretically have altered size-selective sampling characteristics, our results are in line with other studies that performed size-selective sampling of SARS-CoV-2 virus. For instance, Adenaiye et al., analysed SARS-CoV-2 virus in exhaled breath collected from 49 COVID-19 cases (mean days post-onset 3.8 ± 2.1) in an experimental setting and found SARS-CoV-2 RNA in 36% of fine (≤5µm), and 26% of coarse (>5µm) aerosols ^27^. Moreover, other studies using the same CDC-NIOSH bio-sampler methodology as this study, exclusively detected SARS-CoV-2 in the larger ≥4µm and intermediate size fraction 1-4µm in environmental samples collected in rooms of COVID-19 patients in hospitals ^4,5^. Similar observations in size distribution have been reported previously for human influenza virus ^28,29^. These results for different viruses from infected patients indicate that a substantial part of particulates is found in the respirable fraction ^28^. Viral RNA loads and infectious viral RNA loads can differ between patients and are likely influenced by infection status and disease progression. Moreover, the strain-specific viral load and the location of infection in airways influence the particle size distribution and transmission mode to the environment. A different variant, such as Omikron, which is more contagious and is primarily present in the upper respiratory tract, might therefore distribute differently in the environment ^30^.

Out of ten active air samples eligible for culture, we were able to replicate virus from one sample. Only a few studies successfully showed signs of SARS-CoV-2 replication in air samples ^1,8,27^. However, underestimation of infectiousness is a likely consequence of virus inactivation during sample collection ^29^. Current culture techniques may not be optimal for low viral concentrations as in air samples ^31^. Overall, results suggest that virus particulates can cause infection in individuals who inhale these particulates when the infectious dose is sufficiently high.

Literature on the infectious dose of SARS-CoV-2 is scarce. Dabisch et al. reported an infectious dose of 52 TCID50 for a seroconversion response and 256 TCID50 for a fever response based on an inhalation exposure of 10 minutes in non-human primates Macaques ^32^. Others have estimated an infectious dose for infection ranging between single and 1000 virions based on a model combining information on viral mutations obtained through deep sequencing and epidemiology in known infector-infectee pairs ^33–35^. Based on the estimated relationship between E-gene RT-PCR Cq values and cell-cultured SARS-CoV-2 virus loads by Schuijt et al., the air sample which showed replication in our study contained approximately 170 000 viral copies per cubic meter of air ^36^. Despite uncertainties associated with this simple calculation (for instance, assuming similarity in RT-qPCR responses between cell-cultured virus and air samples), the estimated dose may indeed be capable of causing infection. Moreover, our measurements took place during relatively long periods. Environmental levels likely varied considerably over the sampling period. Variation in viral load could not be established over this time span. However, it is unlikely that viral shedding is constant over time. Coughing, for instance, results in higher viral RNA loads over a short time span.

There is an ongoing debate on the airborne transmission route of SARS-CoV-2 and the effect of ventilation on airborne transmission. Greenhalg et al. previously pointed out multiple reasons for airborne transmission as the main route of SARS-CoV-2 ^37^, to which our study provides additional strength. First, our study detected SARS-CoV-2 in abundance in air and on surfaces, including numerous low-touch surfaces such as on top of the wardrobe, which implicates viral dissemination through the air by aerosols. Second, SARS-CoV-2 was primarily found in particle size fractions of 1-4μm and larger than 4 μm, which are known to stay airborne for extended periods of time and thus disseminate potentially over larger distances. Third, we successfully cultured SARS-CoV-2 from an active air sample from particle size >4μm and aerosols have been reported to stay infectious in the air for up to 3 hours ^38^.

Based on our study, ultra-fine particles (<1μm), which can travel further, do not seem to be the key vehicle of SARS-CoV-2 transmission. Although virus contamination was omnipresent in air in infected patient rooms, the vast majority of settling dust and surface swab samples from common areas were negative, suggesting SARS-CoV-2 transmission is more a local phenomenon than widespread. To mitigate (occupational) transmission risks, it is important to investigate the effect of ventilation and air filtration on airborne transmission reduction. To date, only Morris et al. investigated and successfully demonstrated removal of SARS-CoV-2 from air by placing active filtration and sterilisation devices in wards ^5^. Further research on the effect of ventilation and filtration devices is required to draw strong conclusions about the role of ventilation conditions in reducing airborne transmission.

In conclusion, we gained insight into the extent of SARS-CoV-2 presence in air and on surfaces in case of actively shedding patients. Furthermore, the environmental contamination of SARS-CoV-2 and infectious aerosols confirm the potential for transmission via air up to several meters. These insights can contribute to the discussion on airborne transmission and facilitate effective design of prevention strategies such as use of facemasks and optimising ventilation conditions.

## Data Availability

All data produced in the present study are available upon reasonable request to the authors

## Acknowledgements

We thank the patients and healthcare workers for their cooperation and in particular Michelle van Wanrooij and Adrie de Laat from the overarching healthcare organisation Mijzo Waalwijk for their commitment and contribution. We further thank our colleagues Daan Cohen, Calvin Gue, Kees Meliefste, Duco Ottevanger, Santiago Parga, Myrna de Rooij, Peter Scherpenisse and Wouter van der Hoef from Institute Risk Assessment, Lennie Derde and Etienne Sluis from the University Medical Center Utrecht for their contribution in optimalisation of air sampling, laboratorial preparations and sample processing, and Microvida location Amphia Roosendaal and Department of ViroScience of Erasmus MC Rotterdam for further analysis of the samples. Moreover, we thank Dr. Lindsley from National Institute for Occupational Safety and Health Morgentown for the CDC-NIOSCH bio-samplers for their assistance in the pilot study. This study is funded by ZonMw and part of Control of COVID-19 iN Hospitals (COCON) consortium which also involves Rosa van Mansfeld, Karin-Ellen Veldkamp, Andreas Voss and Herman Wunderink.

## Supplement: method and materials

### Outbreak investigation

#### Filtration-based sampler

The filtration-based technique captures inhalable dust - airborne particles and droplets of an aerodynamic size that enter the respiratory tract through mouth and nose. Air is drawn through an inhalable dust sampling head (Conical Inhalable Sampler, JS Holdings, Stevenage, UK) equipped with 37mm diameter 2.0 μm pore-size Teflon filter (Pall incorporated, Ann Arbor, USA) connected with tubing to a Gilian GilAir 5 pump (Sensidyne, St. Petersburg, USA) calibrated at a flow of 3.5 L/min. The sampling head were attached to a pole at 1.5m height as this is average breathing height. The sampling train was set up before the beginning of the measurement and lasted for 6 hours. In each patient room, three inhalable dust samplers were placed, one near the head of the patient, one near the feet of the patient and one near the location often used by healthcare worker (Figure S1).

#### Cyclone-based sampler

The cyclone-based technique allowed for size-selective sampling of dusts and aerosols from the environment. The CDC-NIOSH cyclone-based bioaerosol sampler (NIOSH BC 251, kindly provided to us by William G. Lindsley, NIOSH Morgantown, USA) consists of a sampling body which is equipped with a 15ml conical tube (Greiner Bio-One, Alphen aan de Rijn, Netherlands) to capture aerosols larger than 4 μm in size, a 1.5ml conical tube (Sarstedt BV, Etten-Leur, Netherlands) to capture aerosols with a size ranging between 1-4 μm, and a 37mm diameter filter holder (SKC Incorporated, Eighty Four, USA) equipped with 37mm diameter 2.0 μm pore-size Teflon filter (Pall incorporated, Ann Arbor, USA) to collect particles of 1 μm and smaller. The filter holder was connected with tubing to a Gilian GilAir 5 pump (Sensidyne, St. Petersburg, USA) calibrated at a flow of 3.5 L/min. The 15ml and 1.5ml tube were pre-filled with 2.5ml and 1ml virus transport medium 1 (VTM-1; Erasmus Medical Center (EMC), Rotterdam, The Netherlands), respectively, before start of the measurement (table S1 composition of media). The sampling train was set up before the beginning of the measurement and lasted for 6 hours. In each patient room, one cyclone sampler was placed in conjunction with one of the filtration-based samplers close to the feet of the patient. After sampling filter holders were detached, packed in a Minigrip ™ bag. The 15ml and 1.5ml tubes were detached from the sampling body, 1 ml Opti-MEM™ (Gibco, UK) and 1 ml of VTM-1/Opti-MEM ™ mixture, respectively, was added to the tubes immediately after sampling.

#### Impingement-based sampler

Air sampling through impingement was conducted by means of a 5ml BioSampler (SKC Inc, Eighty Four, USA) positioned at 1.5m height attached to a pole. An airflow of 12.5 L/min through the BioSampler was established by connecting the outlet of the sampler to an inhouse designed pump unit. The impinger was filled with 4ml of VTM-1 used for air sampling (see above) prior to the beginning of the measurement. The measurement lasted 1 hour close to the patient’s head, during which evaporation losses of VTM was replaced by adding every 15 minutes 2 ml of VTM-1. After sampling, remaining VTM fluid was transferred to a 15ml tube (Greiner BioOne, Etten-Leur, Netherlands) and 2 ml of Opti-MEM ™ was added.

#### Settling dust sampler

Settling dust, passive air, samples were collected by using Electrostatic Dust Collectors (EDCs) (Noss et al., 2008), which were placed in each included patient room and corresponding hallway, common living room and nurse office of the ward. EDCs were placed in holders pinned to the ceiling in the middle of the space. After 2-4 weeks of sampling EDCs were picked up and packed in a Minigrip™ bag for transportation.

#### Collection of surface samples

In each of the above areas: each included patient room, hallway, living room and nursing office, ten swab samples from surfaces were collected. Sampling locations included high-touch surfaces like door handle and tabletop and low-touch surfaces like top surface of cabinets. To standardize swabbing of surfaces, disposable plastic grids of 10 cm^2^ were used; when it was not possible to use the grid this was noted. Dry swabs (Medical Wire Dry Swabs, MW370, Corsham, UK) were used, which were placed in 2ml viral transport medium 2 (VTM-2; Erasmus Medical Center (EMC), Rotterdam, The Netherlands) in 5ml tubes directly after swabbing (for composition of media see table S2).

#### Patient characteristics

Oropharyngeal swabs (OPS) was collected from patients during sample collection. Dry swabs (Medical Wire Dry Swabs, 111598, Milano, Italy) were used, which were placed in 2ml VTM-2 in 5ml tubes directly after swabbing.

### Laboratory analysis

After collection, all samples, except for EDCs, were stored at 4 °C in electric transport cooling box and transported to the lab. At the lab, all samples, except for EDCs, were placed in 4°C storage until further processing the next day. All handlings were performed under BSL2+ conditions. In short, filters were removed from the filter holders and transferred to 5ml screw-top tubes (Eppendorf, Nijmegen, Netherlands) and 2ml of VTM-1 used for air sampling was added, next tubes were vortexed for 5 minutes using a vortex adaptor. Tubes containing VTM and Opti-MEM™ mixture from NIOSH and impingement-based samplers and OPS were vortexed for 15 seconds, and tubes containing the surface swab samples were vortexed for 1 min prior to further handling. After vortexing 600 μl was transferred and added to a tube containing 600 μl of MagNA Pure 96 External Lysis Buffer (Roche Diagnostics, Almere the Netherlands), followed by 15 seconds of vortexing. Aliquots with remaining VTM from NIOSH, impingement, surface swab and OPS were stored frozen at -80 °C for culturing. Aliquots with VTM/lysis buffer samples were stored frozen at -80 °C for PCR.

#### Settling dust sampler

EDCs were transferred to tube containing 10 ml VTM-2. EDCs were tamped down with disposable pipette, followed by repeating twice 15 seconds of vortexing and soaked for 15 minutes and ended with 15 minutes of vortexing. EDCs were tamped down with disposable pipette and 600 ul sample was transferred and added to a tube containing 600 ul MagNA Pure 96 External Lysis Buffer (Roche Diagnostics, Almere the Netherlands) and followed by 15 seconds of vortexing. VTM/lysis buffer samples were stored frozen at -80 °C.

#### Real time quantitative RT-PCR

Samples were transported on dry ice to Microvida Laboratory for Medical Microbiology, Bravis Hospital, Roosendaal, The Netherlands. Presence of SARS-CoV-2 RNA was tested using a SARS-CoV-2 RNA RT-qPCR, targeting the E gene and CoV-2 RdRP-gene of SARS-CoV-2. The cobas® SARS-CoV-2 Test (Roche Diagnostics, Basel, Switzerland) was performed on the samples with lysis buffer using the cobas® 6800 Systems (Roche Diagnostics). Samples were positive if the threshold was below Ct-value 40.

### SARS-CoV-2 Whole genome sequencing (WGS)

Samples with RT-PCR RdRp Ct-values <31 whole genome sequencing of the primary clinical specimen was performed by Microvida to determine the SARS-CoV-2 variant. In short, total nucleic acids were extracted using the QIAsymphony DSP virus pathogen midi kit and pathogen complex 400 protocol of the QIAsymphony Sample Processing system (Qiagen, Germany), with an input volume of 400 μL and output volume of 110 μL. cDNA was synthesized using LunaScript® RT SuperMix Kit (New Engeland Biolabs, USA) and library preparation was performed using EasySeqTM RC-PCR SARS-CoV-2 Whole Genome Sequencing kit (Nimagen, The Netherlands) according to manufacturer’s instructions. Subsequent next generation sequencing (NGS) of 2×150cycles paired end reads was performed on a Miseq (Illumina, The Netherlands) using MiSeq Reagent Micro Kit v2 according to manufacturer’s instructions. Data analyses was performed with an in-house workflow using CLCbio Genomic Workbench v21 (Qiagen, Germany), including (a.o.) read-trimming, NC_045512.2 (NCBI genbank) reference-based assembly, local re-alignment and variant detection algorithms. The consensus genome was extracted and positions with a coverage less than 10 reads were replaced with N. The sequences were manually curated. Genomes with >70% genome coverage were included for lineage assignment using Pangolin (https://pangolin.cog-uk.io/)^39^ and Nextclade Web (https://clades.nextstrain.org/) (Hadfield et al., 2018)). Genomes with >90% genome coverage were uploaded to GISAID (https://www.gisaid.org/)^40^, with accession IDs EPI_ISL_3047866, EPI_ISL_3047867, EPI_ISL_2259188, EPI_ISL_2259136 and EPI_ISL_2259122 and phylogenetic and analysis performed using MAFFT alignment software (Galaxy Version 7.221.3, FFT-NS method) ^41^ and phylogenomic software IQ-TREE 2.1.3 (Galaxy Version 1.5.5.3)^42–44^, using ModelFinder (with best predicted method GTR+F+I), ultrafast bootstrapping (1000 replicates) and maximum likelihood tree reconstruction. Phylogenetic tree was visualized in CLCbio Genomic Workbench v20.

#### Virus culture

Positive tested samples with RT-qPCR from the OPS, cyclone-based, impingement-based and surface swabs were transported on dry ice, using the duplex aliquot without lysis buffer for culturing at Erasmus University Medical Centre laboratory, Rotterdam, the Netherlands. Culturing was performed on Vero cells, clone 118 at 37°C, and 5% CO2 and was completed after 7 days. If virus-induced cytopathic effect was observed, immunofluorescent detection of nucleocapsid proteins was performed to confirm the presence of SARS-CoV-2^16^.

#### Modification in sample collection

After the first outbreak investigation outbreak A, an optimised protocol was implemented. For the first outbreak investigation VTM-2 was used as pre-fill in the tubes for the cyclone-based, impingement-based and re-fill for impingement sampling in instead of VTM-2. After sampling the impingement-based sampler VTM fluid was transferred to a 15 ml tube (Greiner BioOne, Etten-Leur, Netherlands) containing 1,5 ml Fetal Bovine Serum (FSB) (40 v/v%, Greiner Bio-One, Etten-Leur, Netherlands). Instead of adding Opti-MEM™ mixture or VTM/Opti-MEM™ mixture after sampling to the cyclone-based samples, the day after during processing only VTM-2 was added up to the original amount prior to sample collection. Sampling and processing of surface swabs, OPS and EDCs stayed identical.

**Figure S1:**
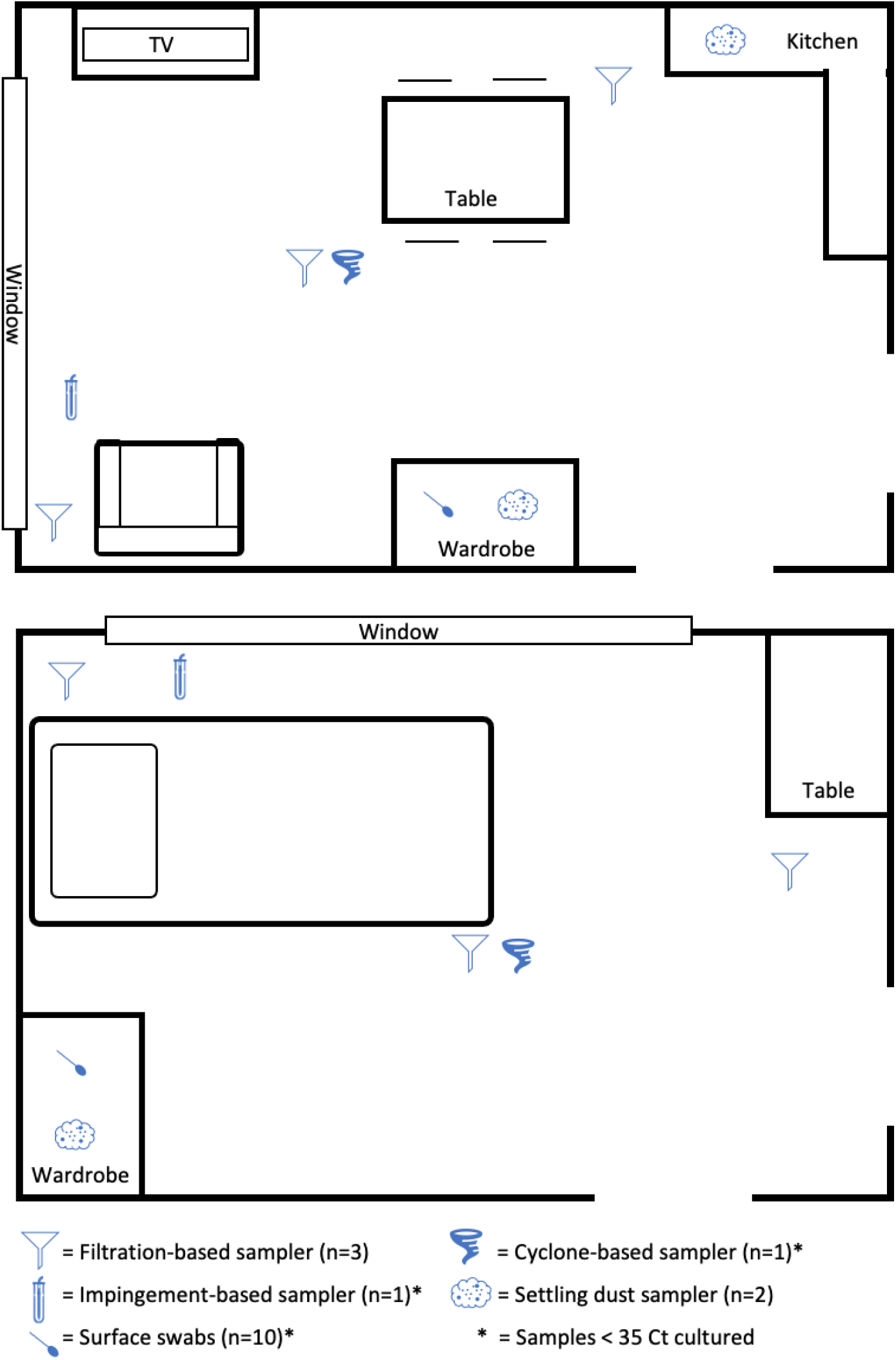
Illustration of sample collection in patient room. The first figure shows data collection in the living room of a mobile patient sitting in a chair. The second figure illustrates data collection in the sleeping room of a bedridden patient.

**Table S1:**
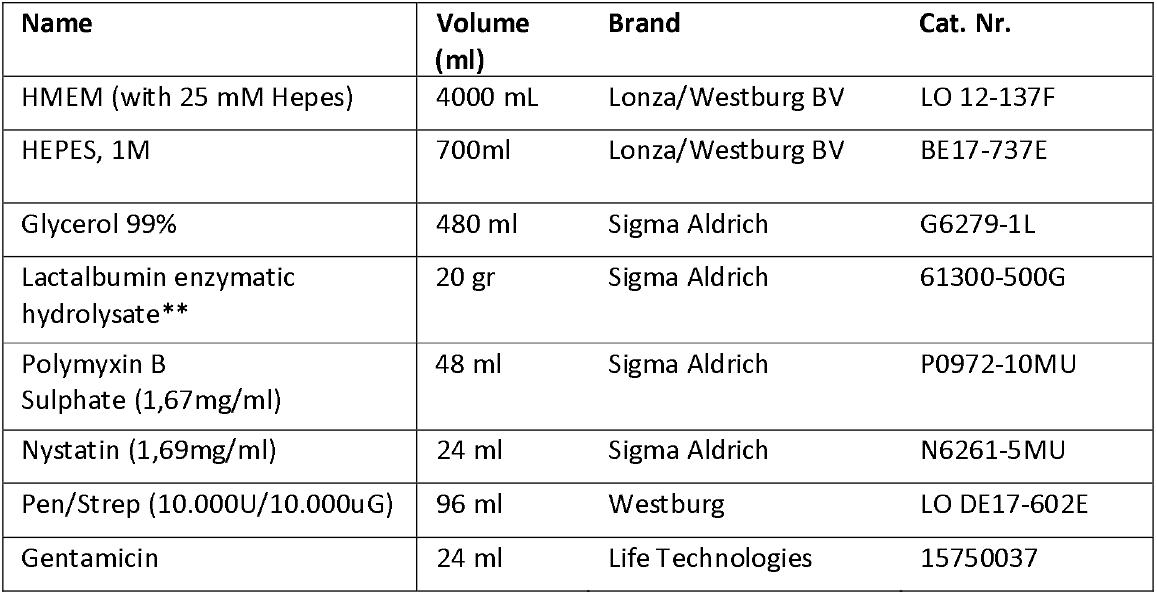
VTM-1 composition.

**Table S2:**
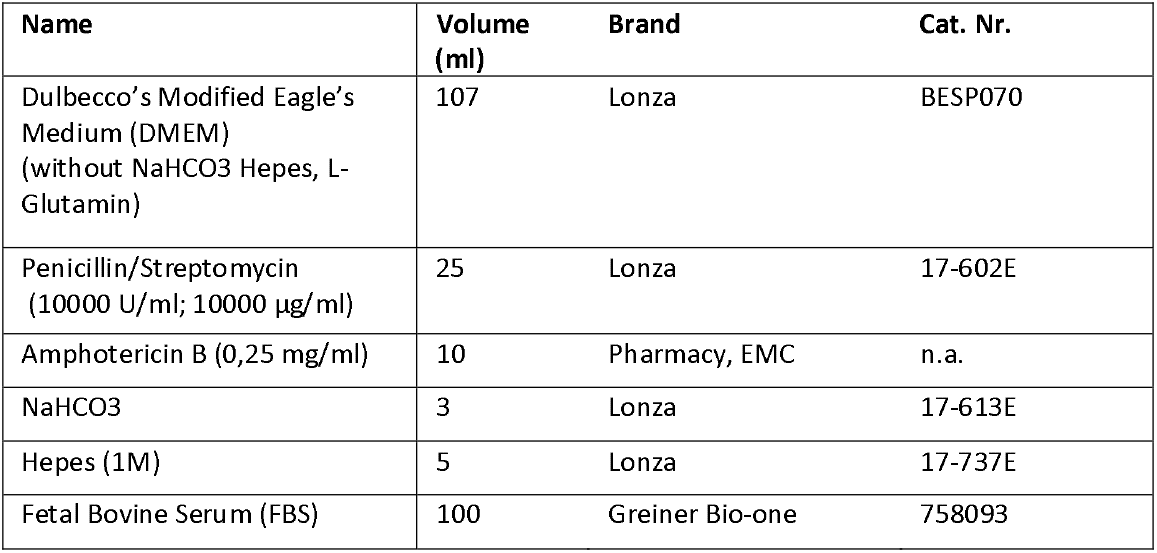
VTM-2 composition.

## Supplement: Results

**Table S3:**
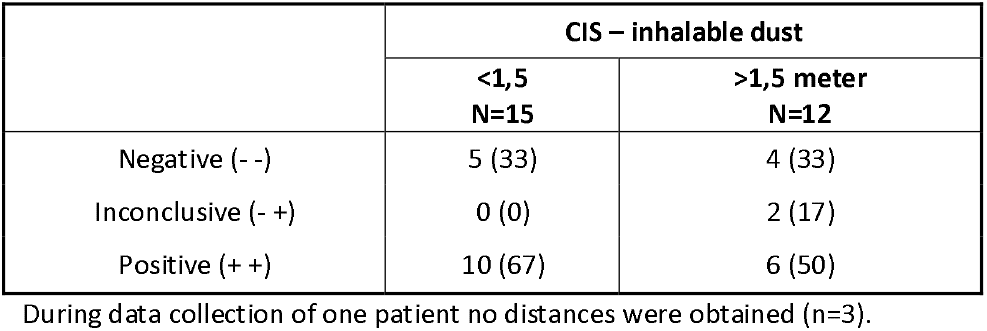
SARS-CoV-2 PCR results in inhalable dust samples collected in rooms of patients with positive OS stratified by distance to the patient.

**Table S4:** SARS-CoV-2 PCR results in inhalable dust samples cyclone-based samples from patients with positive OS.

|  | CIS -<br>inhalable dust | CDC-NIOSH bioaerosol sampler -<br>cyclone-based | Distance (cm) |
| --- | --- | --- | --- |
| Patient room 10 | ++ | ++ | NA |
| Patient room 12 | ++ | ++ | 160 |
| Patient room 13 | ++ | ++ | 100 |
| Patient room 15 | -- | -- | 210 |
| Patient room 16 | ++ | ++ | 85 |
| Patient room 17 | ++ | ++ | 74 |
| Patient room 18 | ++ | ++ | 140 |
| Patient room 20 | ++ | ++ | 135 |
| Patient room 22 | ++ | -- | 100 |
| Patient room 25<sup>V</sup> | -- | - + | 130 |

<sup>V</sup> = fully vaccinated patient

**Table S5:** SARS-CoV-2 PCR results surface swab samples from patients with positive OS.

|  | High-touch | Low-touch |
| --- | --- | --- |
| Negative (- -) | 33 (60) | 19 (43) |
| Inconclusive (- +) | 1 (2) | 3 (7) |
| Positive (+ +) | 21 (38) | 22 (50) |

**Table S6:** SARS-CoV-2 PCR results surface swab samples from common areas.

|  | High-touch | Low-touch |
| --- | --- | --- |
| Negative (- -) | 94 (98,9) | 98 (96) |
| Inconclusive (- +) | 0 (0) | 0 (0) |
| Positive (+ +) | 1 (1,1) | 4 (4) |

**Table S7:**
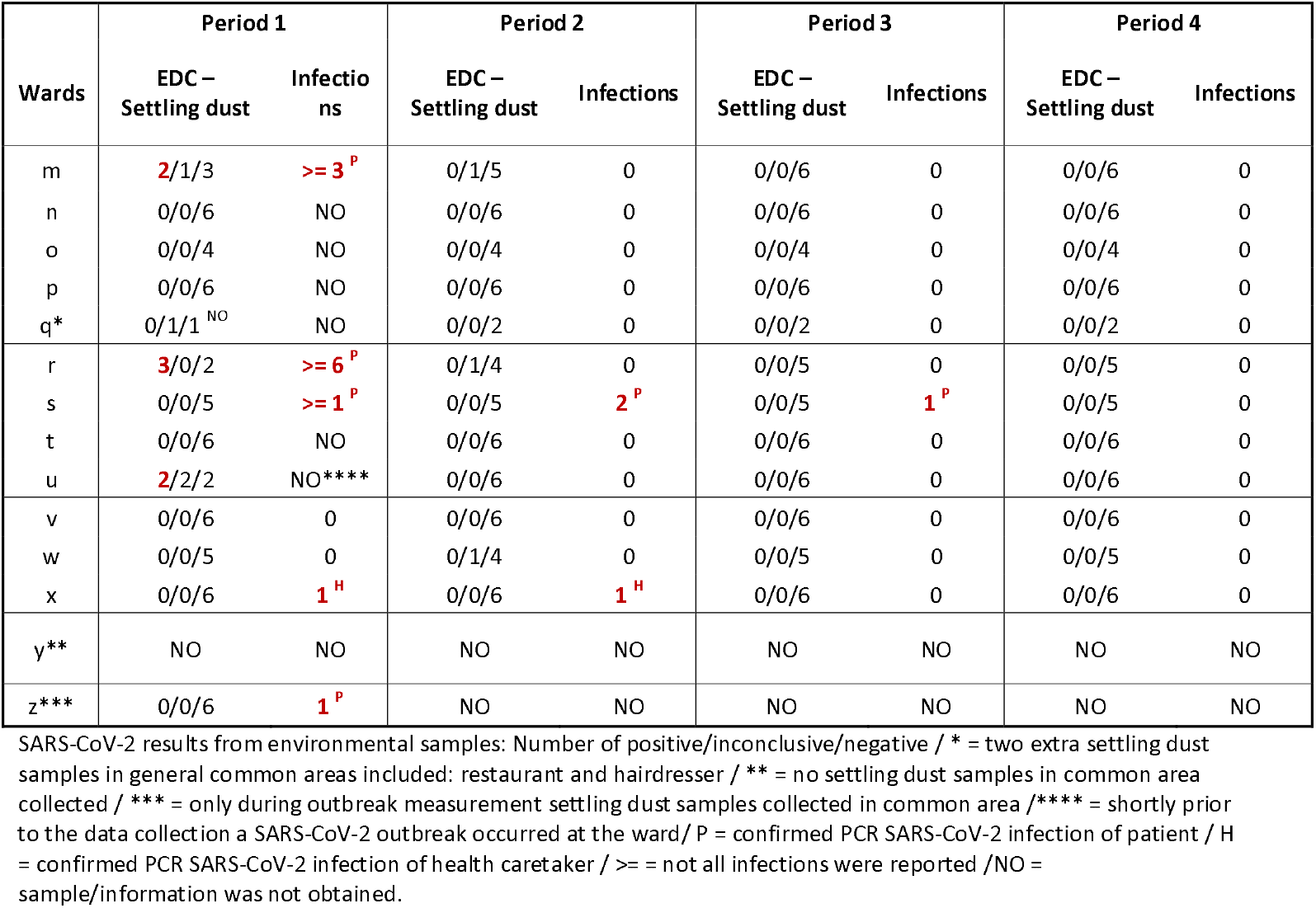
SARS-CoV-2 PCR in environmental samples collected at nursing homes wards and reported SARS-CoV-2 infections from corresponding nursing homes.

| Wards | Period 1 |  | Period 2 |  | Period 3 |  | Period 4 |  |
| --- | --- | --- | --- | --- | --- | --- | --- | --- |
|  | EDC –<br>Settling dust | Infections | EDC –<br>Settling dust | Infections | EDC –<br>Settling dust | Infections | EDC –<br>Settling dust | Infections |
| m | 2/1/3 | >= 3<sup>P</sup> | 0/1/5 | 0 | 0/0/6 | 0 | 0/0/6 | 0 |
| n | 0/0/6 | NO | 0/0/6 | 0 | 0/0/6 | 0 | 0/0/6 | 0 |
| o | 0/0/4 | NO | 0/0/4 | 0 | 0/0/4 | 0 | 0/0/4 | 0 |
| p | 0/0/6 | NO | 0/0/6 | 0 | 0/0/6 | 0 | 0/0/6 | 0 |
| q* | 0/1/1<sup>NO</sup> | NO | 0/0/2 | 0 | 0/0/2 | 0 | 0/0/2 | 0 |
| r | 3/0/2 | >= 6<sup>P</sup> | 0/1/4 | 0 | 0/0/5 | 0 | 0/0/5 | 0 |
| s | 0/0/5 | >= 1<sup>P</sup> | 0/0/5 | 2<sup>P</sup> | 0/0/5 | 1<sup>P</sup> | 0/0/5 | 0 |
| t | 0/0/6 | NO | 0/0/6 | 0 | 0/0/6 | 0 | 0/0/6 | 0 |
| u | 2/2/2 | NO**** | 0/0/6 | 0 | 0/0/6 | 0 | 0/0/6 | 0 |
| v | 0/0/6 | 0 | 0/0/6 | 0 | 0/0/6 | 0 | 0/0/6 | 0 |
| w | 0/0/5 | 0 | 0/1/4 | 0 | 0/0/5 | 0 | 0/0/5 | 0 |
| x | 0/0/6 | 1<sup>H</sup> | 0/0/6 | 1<sup>H</sup> | 0/0/6 | 0 | 0/0/6 | 0 |
| y** | NO | NO | NO | NO | NO | NO | NO | NO |
| z*** | 0/0/6 | 1<sup>P</sup> | NO | NO | NO | NO | NO | NO |

SARS-CoV-2 results from environmental samples: Number of positive/inconclusive/negative / \* = two extra settling dust samples in general common areas included: restaurant and hairdresser / \*\* = no settling dust samples in common area collected / \*\*\* = only during outbreak measurement settling dust samples collected in common area /\*\*\*\* = shortly prior to the data collection a SARS-CoV-2 outbreak occurred at the ward/ P = confirmed PCR SARS-CoV-2 infection of patient / H = confirmed PCR SARS-CoV-2 infection of health caretaker / >= = not all infections were reported /NO = sample/information was not obtained.

**Figure S2:**
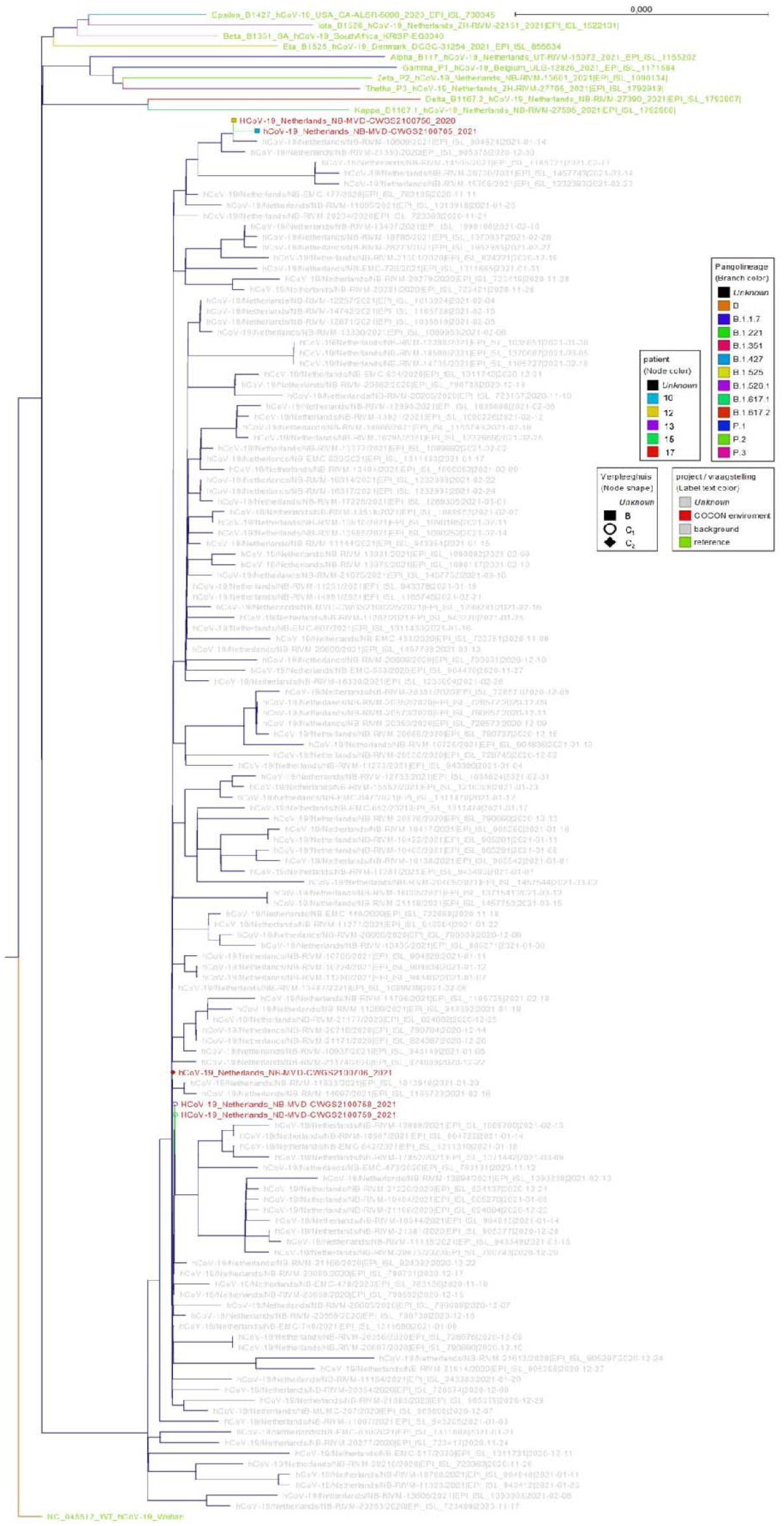
A phylogenetic analysis of samples with minimum >90% reference coverage.

**Table S8:**
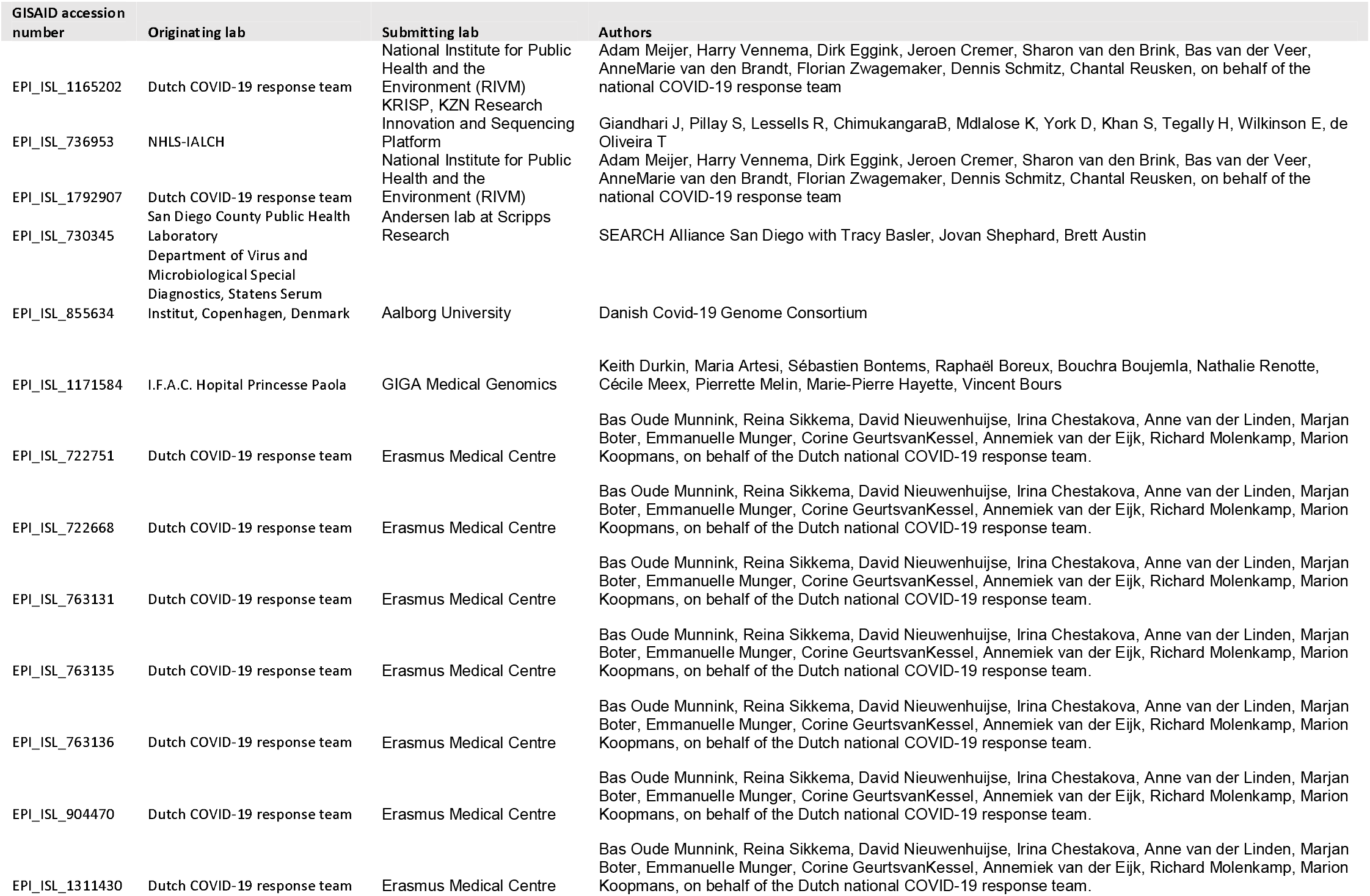
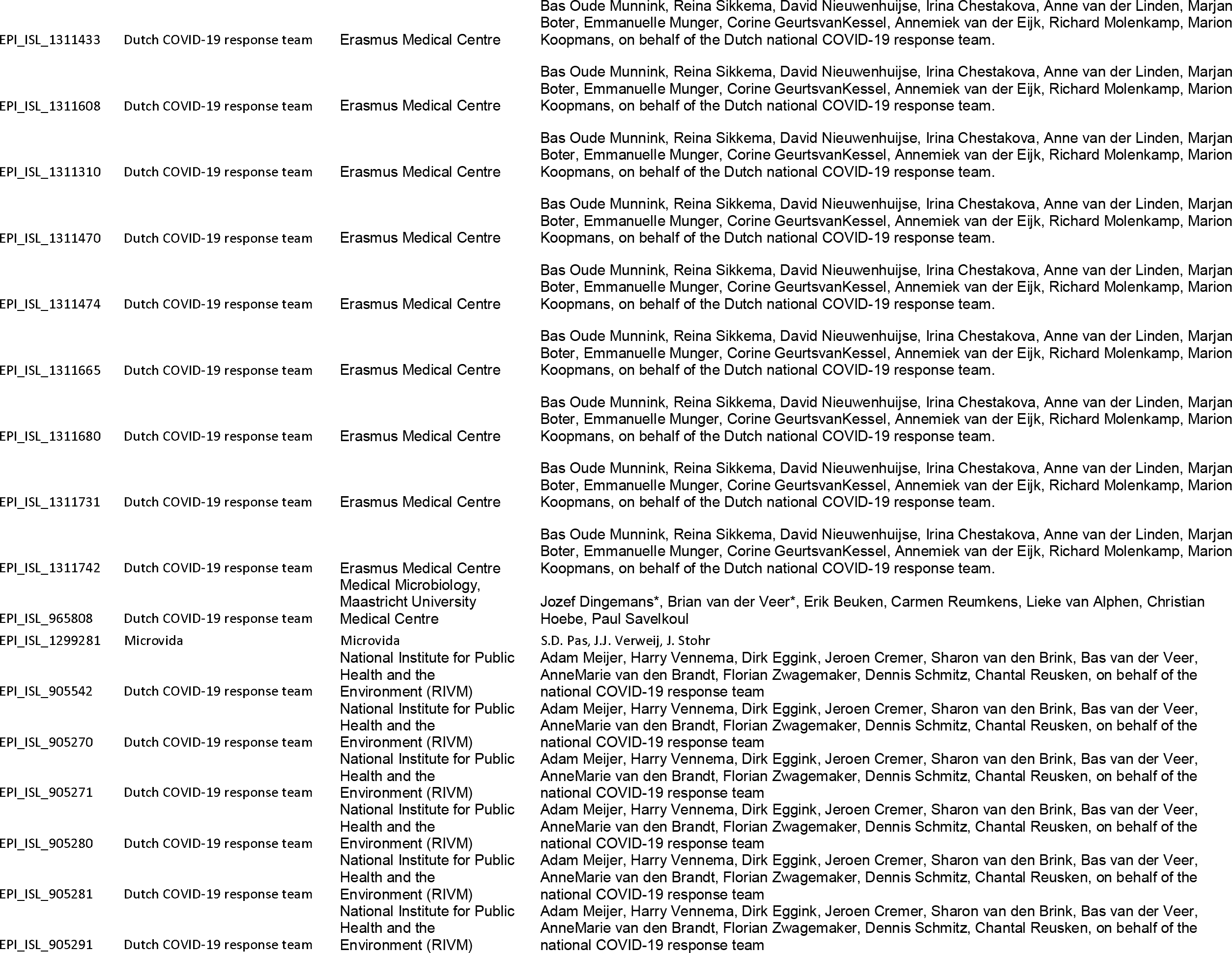
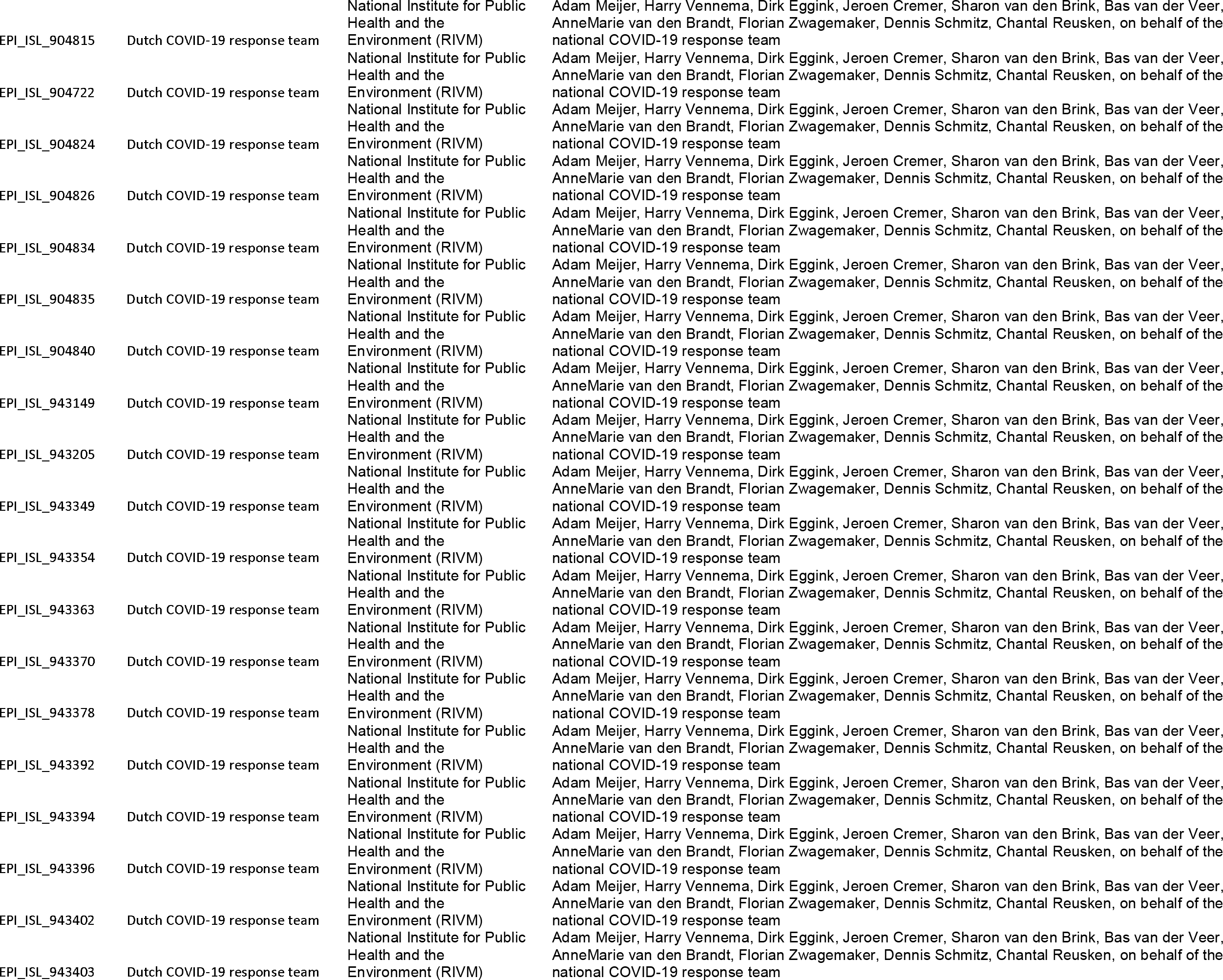
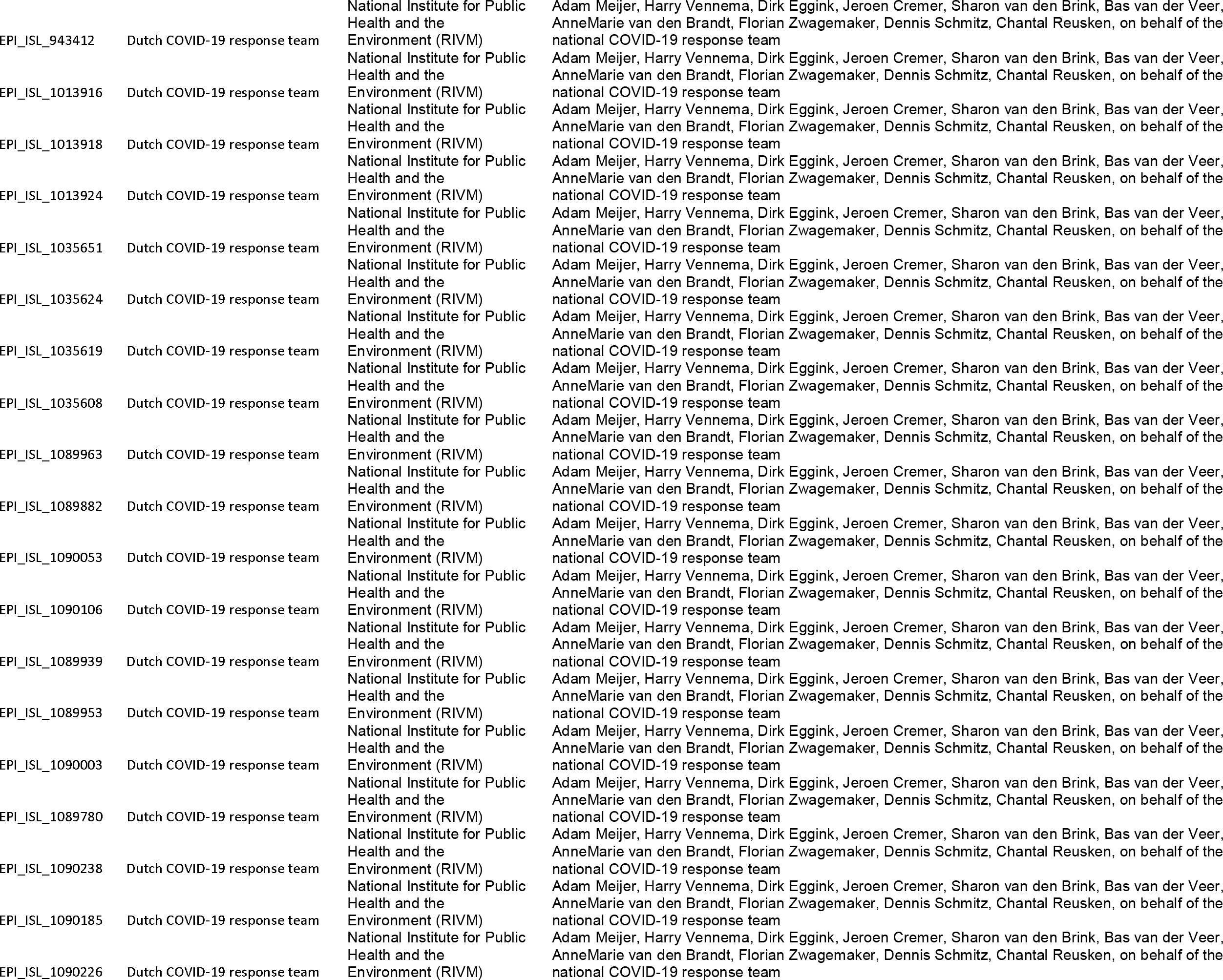
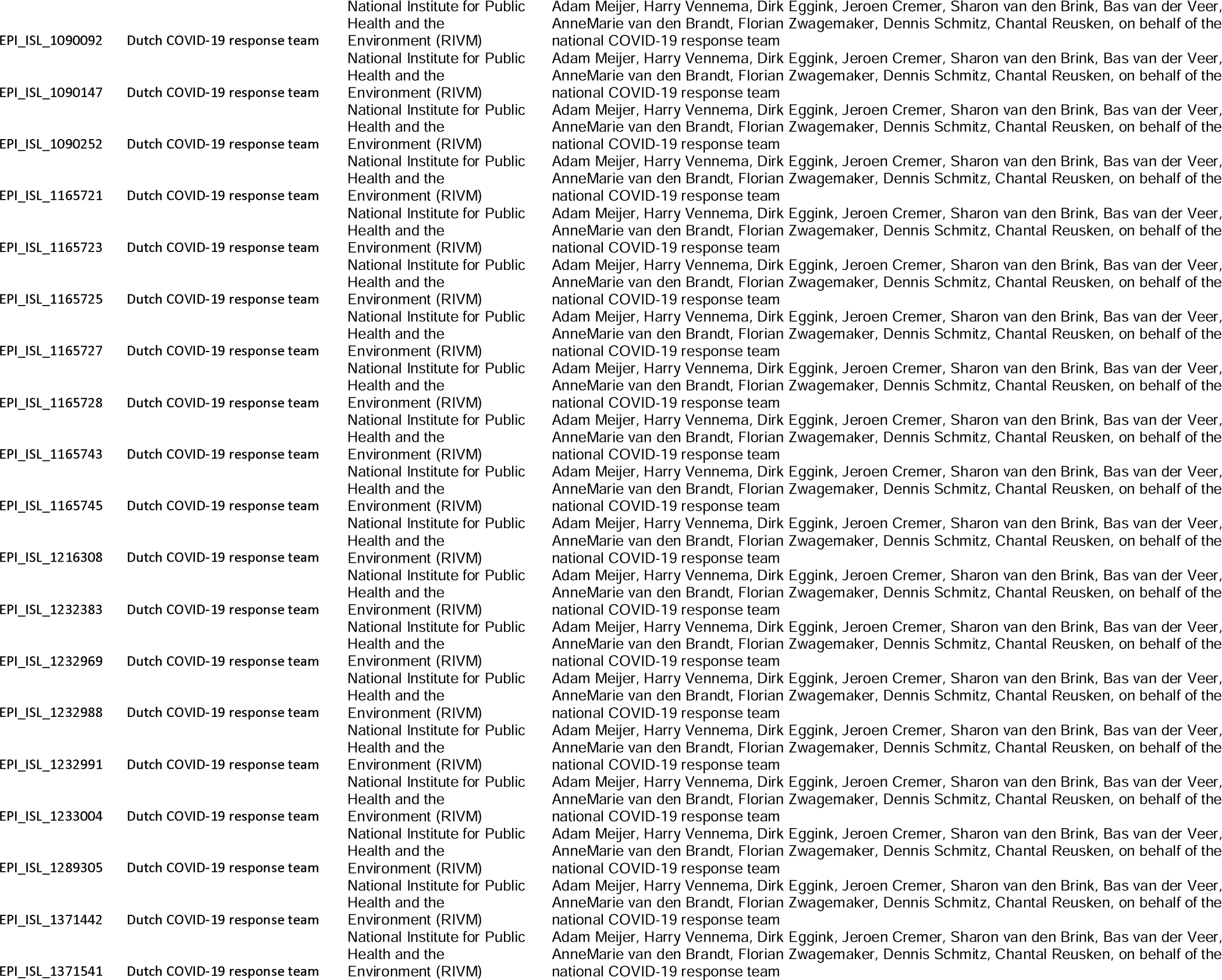
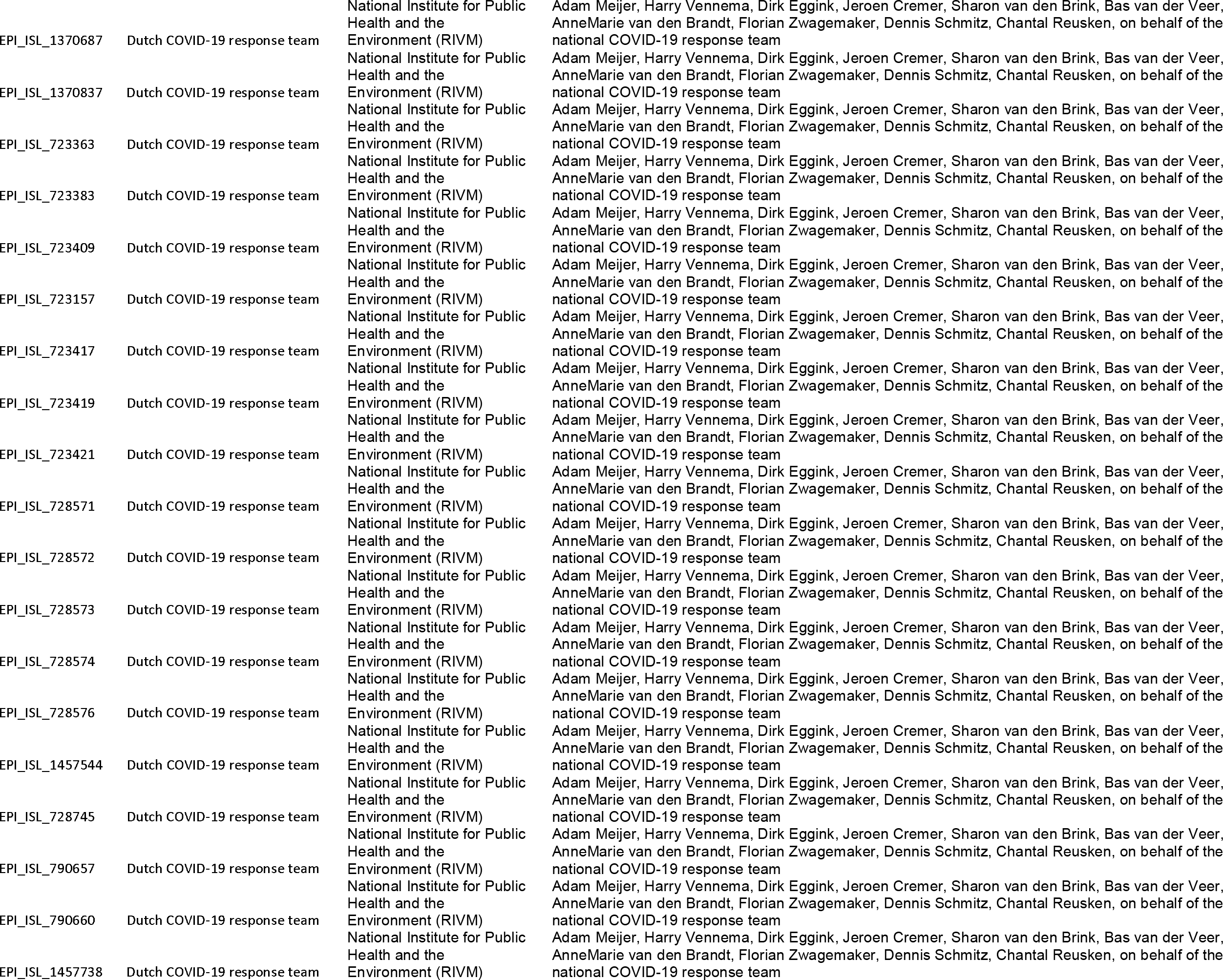
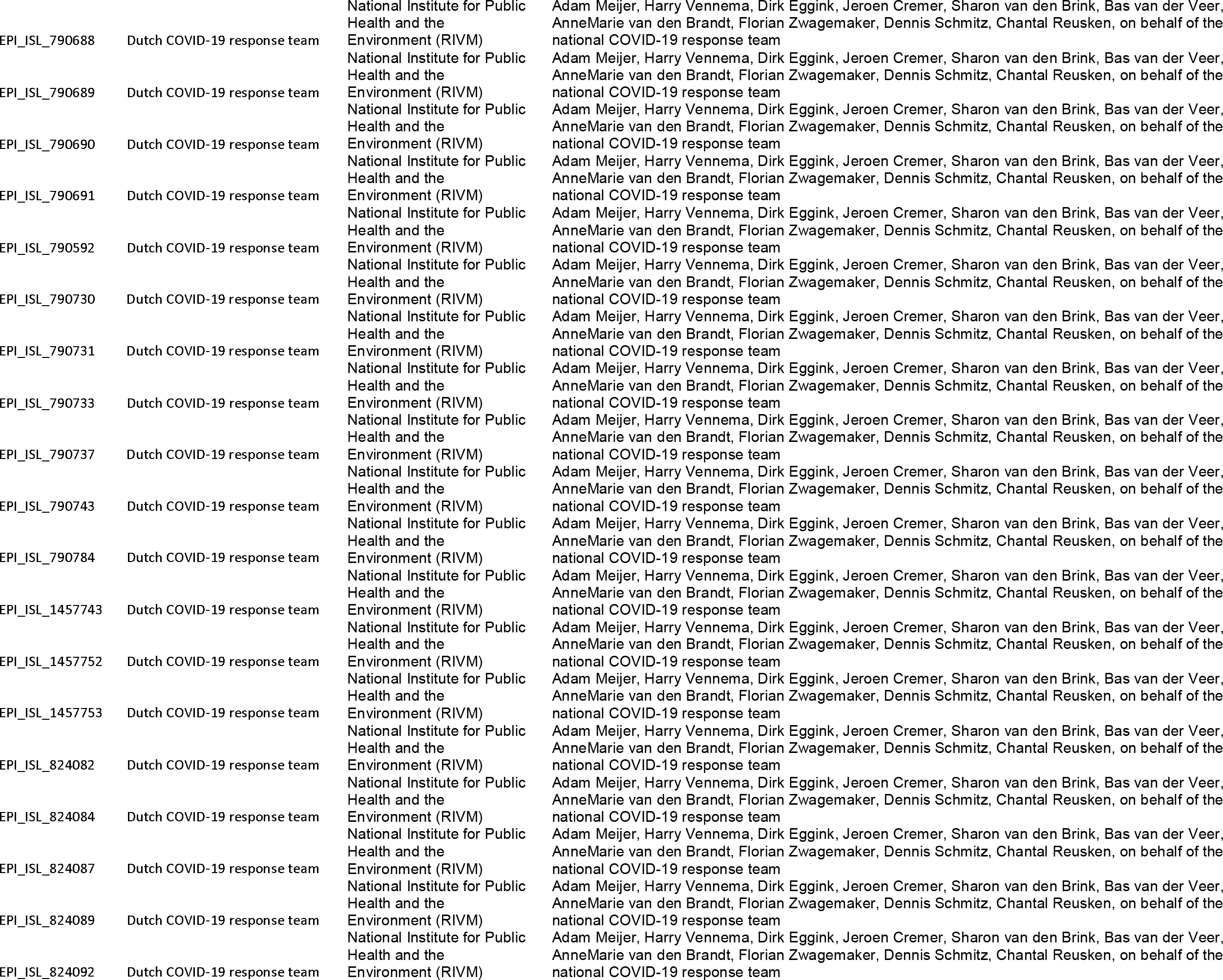
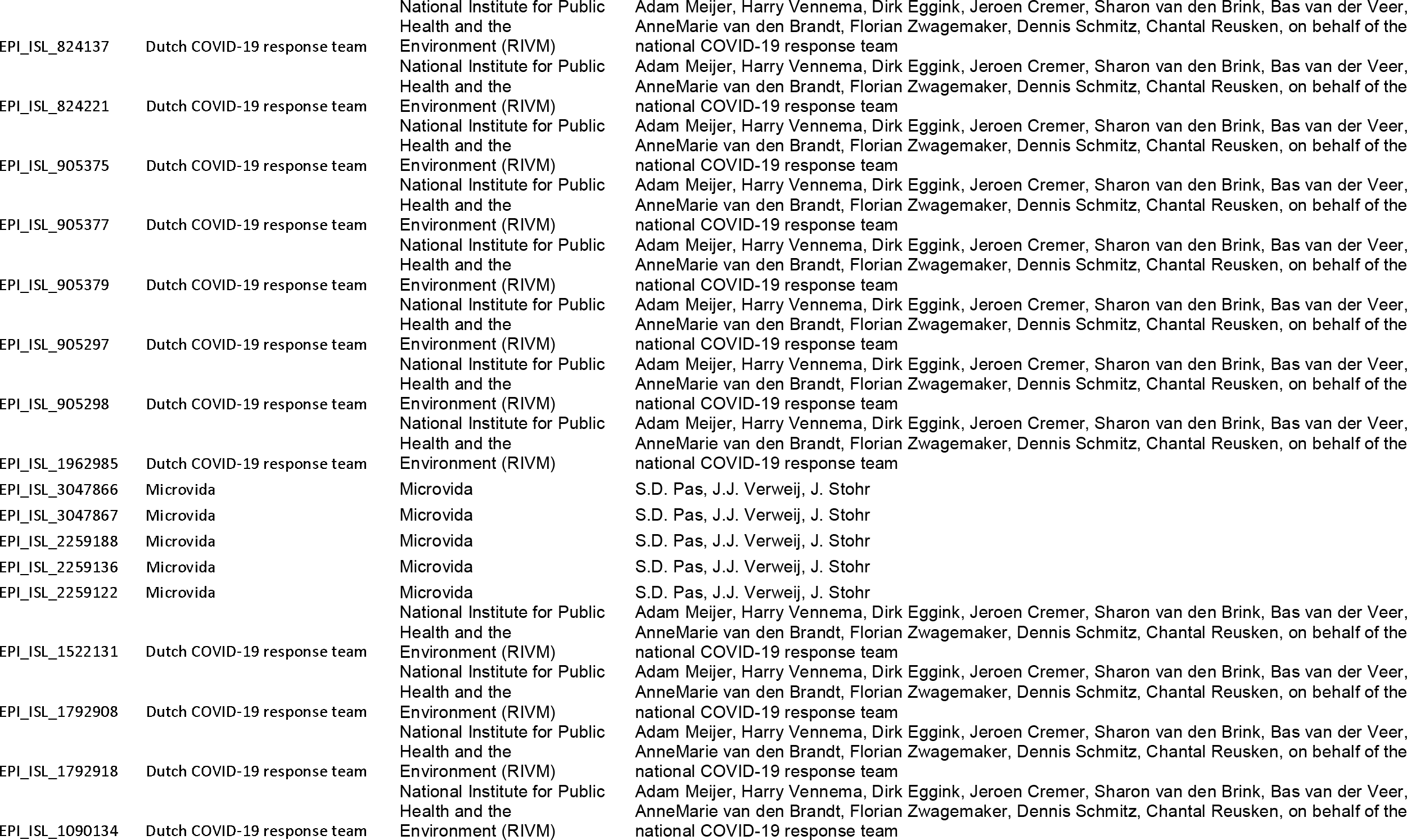
Acknowledgement table Whole Genome Sequencing. We gratefully acknowledge the following Authors from the Originating laboratories responsible for obtaining the specimens, as well as the Submitting laboratories where the genome data were generated and shared via GISAID, on which this research is based. All Submitters of data may be contacted directly via www.gisaid.org. Authors are sorted alphabetically.

## Notes

### Competing Interest Statement

The authors have declared no competing interest.

### Author Declarations

No medical ethical approval was needed for this study as evaluated by the Medical Research Ethics Committee of University Medical Centre Utrecht; a declaration of non-compliance with the scope of the Dutch Medical Research Involving Human Subject Act was obtained.

